# Signatures of transmission in within-host *M. tuberculosis* variation

**DOI:** 10.1101/2023.12.28.23300451

**Authors:** Katharine S. Walter, Ted Cohen, Barun Mathema, Caroline Colijn, Benjamin Sobkowiak, Iñaki Comas, Galo A. Goig, Julio Croda, Jason R. Andrews

## Abstract

**Background:** Because *M. tuberculosis* evolves slowly, transmission clusters often contain multiple individuals with identical consensus genomes, making it difficult to reconstruct transmission chains. Finding additional sources of shared *M. tuberculosis* variation could help overcome this problem. Previous studies have reported *M. tuberculosis* diversity within infected individuals; however, whether within-host variation improves transmission inferences remains unclear.

**Methods:** To evaluate the transmission information present in within-host *M. tuberculosis* variation, we re-analyzed publicly available sequence data from three household transmission studies, using household membership as a proxy for transmission linkage between donor-recipient pairs.

**Findings:** We found moderate levels of minority variation present in *M. tuberculosis* sequence data from cultured isolates that varied significantly across studies (mean: 6, 7, and 170 minority variants above a 1% minor allele frequency threshold, outside of PE/PPE genes). Isolates from household members shared more minority variants than did isolates from unlinked individuals in the three studies (mean 98 shared minority variants vs. 10; 0.8 vs. 0.2, and 0.7 vs. 0.2, respectively). Shared within-host variation was significantly associated with household membership (OR: 1.51 [1.30,1.71], for one standard deviation increase in shared minority variants). Models that included shared within-host variation improved the accuracy of predicting household membership in all three studies as compared to models without within-host variation (AUC: 0.95 *versus* 0.92, 0.99 *versus* 0.95, and 0.93 *versus* 0.91).

**Interpretation:** Within-host *M. tuberculosis* variation persists through culture and could enhance the resolution of transmission inferences. The substantial differences in minority variation recovered across studies highlights the need to optimize approaches to recover and incorporate within-host variation into automated phylogenetic and transmission inference.

**Funding:** NIAID: 5K01AI173385

## Introduction

Reducing the global burden of tuberculosis urgently requires reducing the number of incident *M. tuberculosis* infections. Yet the long and variable latency period of TB infection makes it challenging to identify sources of transmission and thus intervene. Genomic epidemiology approaches have been powerfully applied to characterize *M. tuberculosis* global phylogenetic structure, migration and gene flow, patterns of antibiotic resistance, transmission linkages. Yet transmission inference approaches have often failed to identify the majority of transmission linkages in high-incidence settings^1–6^. Further, while previous studies have identified heterogeneity in the number of secondary cases generated by infectious individuals^7^ and risk factors for onwards transmission^8,9^, these are often difficult to generalize. Many critical questions, including the contribution of asymptomatic individuals to transmission, remain unanswered. Novel, accessible approaches to reconstruct high-resolution transmission patterns are urgently needed so that public health programs can identify environments driving transmission and risk factors for onwards transmission.

Commonly used approaches for *M. tuberculosis* transmission inference use single consensus genomes, representing the sequence of the most frequent alleles, from infected individuals. Closely related pathogen genomes are predicted to be more closely linked in transmission chains. For example, *M. tuberculosis* consensus sequences within a given genetic distance^10–13^ are considered clustered and potentially epidemiologically linked. However, *M. tuberculosis* evolves at a relatively slow rate^14^. The result is that there may be limited diversity in outbreaks. Indeed, several genomic epidemiology studies reported that multiple individuals harbored identical *M. tuberculosis* genomes^13,15–17^, making it difficult to reconstruct who infected whom. This highlights a need to recover more informative variation from pathogen genomes. This challenge is not unique to *M. tuberculosis*—COVID-19 outbreak investigations frequently generate large numbers of identical genomes^18^, indicating a broad need for higher-resolution pathogen genomics approaches.

Population-level bacterial diversity within an individual, or within-host heterogeneity, can be attributed to mixed infections, infections with more than one distinct *M. tuberculosis* genotype, or *de novo* evolution, mutations that are introduced over the course of an individual’s infection^19^. Previous research has found that a substantial proportion (10-20%)^19^ of infected individuals harbor mixed infections with genetically diverse populations of *M. tuberculosis*^19–23^. A portion of within-host heterogeneity is likely transmitted onwards^24,25^ and therefore, within-host diversity captures potentially valuable epidemiological information about transmission history^25,26^. Complex infections are also important clinically. Within-host heterogeneity is associated with poor treatment outcomes^20,27^ and hetero-resistance, the presence of both resistant and susceptible alleles within a single infection, reduces the accuracy of diagnostics for antibiotic resistance^27^.

Despite the evidence that within-host *M. tuberculosis* variation is common, there are many open questions about whether shared within-host variation is a predictor of transmission linkage and, more practically, how to recover this level of variation and incorporate it into transmission inferences. Currently, *M. tuberculosis* is most frequently cultured from sputum samples and sequenced with short reads to generate a single consensus sequence^28^. This limits the variation recovered because (a) culture imposes a severe bottleneck^29–31;^ (b) within-host variation, including mixed infections, are often excluded, in part due to a lack of validated methodological approaches for accurate recovery of such variation^25,31^; and (c) repetitive genomic regions, including the PE/PPE gene families, among the most variant-rich and potentially informative regions of the genome, are excluded^32–34^.

Recent work has demonstrated that pathogen enrichment approaches—through either host DNA depletion or pathogen DNA enrichment—can allow *M. tuberculosis* to be sequenced directly from clinical samples, bypassing the need for culture^23,35–39^. But there have not been consistent findings about whether culture-free approaches improve the detection of within-host variation.

*M. tuberculosis* transmission is never directly observed, making it difficult to assess the performance of genomic methods in identifying true transmission pairs. We therefore tested whether household members—as a proxy for epidemiologically linked individuals—shared more minority variants than did unlinked individuals, and whether minority variation could enhance transmission inferences by re-analyzing previously published household transmission studies.

Here, we study household members as a gold standard, the best available proxy for transmission pairs, to test whether shared minority *M. tuberculosis* variation may augment fixed genomic differences in reconstructing epidemiological linkages. In practice, such as in routine population-wide genomic *M. tuberculosis* sequencing by public health laboratories, epidemiological linkages are frequently unknown. Whether a signal of shared minority variation exists in gold standard transmission pairs can then indicate whether shared minority variation might contribute to resolving such unobserved transmission linkages in population-wide genomic data.

## Methods

### Sequence and epidemiological data

We accessed publicly available data from 3 household transmission studies for which both raw sequence data and epidemiological linkages were publicly available: Colangeli et al. (2020)^40^, Vitória, Brazil; Guthrie et al. (2018)^41^, British Columbia, Canada; and Walker et al. 2014, Oxfordshire, England^11^ (Table 1). Sequence data was available from the Sequence Read Archive (PRJNA475130, PRJNA413593, and PRJNA549270). Colangeli et al. cultured sputa on Lowenstein-Jensen (LJ) slants, plated cultures on Middlebrook 7H10 agar, and then scraped three loops of culture for DNA extraction^40^. Both Guthrie et. al and Walker et al. re-cultured frozen isolates on MGIT liquid medium or LJ slants^11,41^. We accessed information on household pairs from published phylogenies in the Colangeli et al. and Guthrie et al. papers. For the Walker et al. paper, household linkages were available in the data supplement.

**Table 1.**
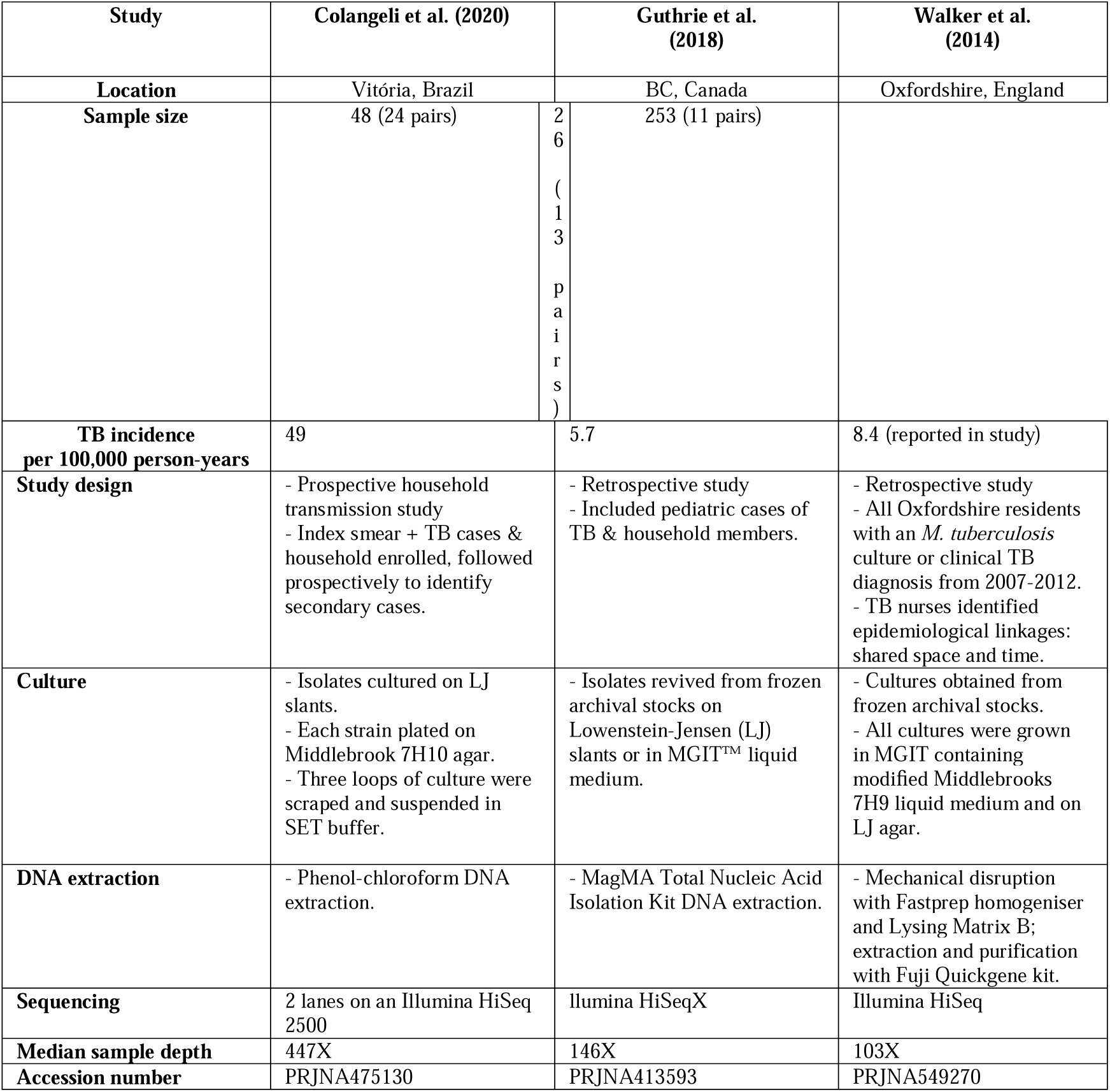
*M. tuberculosis* household transmission study characteristics. TB incidence per 100,000 is from the World Health Organization 2022 Country Profiles unless otherwise noted.

### Bioinformatic analysis

We processed raw sequence data with a previously described variant identification pipeline available on GitHub (https://github.com/ksw9/mtb-call2).^4042^ We previously conducted a variant identification experiment to compare commonly used mapping and variant calling algorithms in *M. tuberculosis* genomic epidemiology^32^. We found that the combination of the *bwa*^43^ mapping algorithm and *GATK*^44,45^ variant caller routinely minimizes false positive variant calls with minimal cost to sensitivity as compared to other tool combinations^46^, especially when the PE/PPE genes are excluded. We therefore used this combination of tools in our pipeline.

Briefly, we trimmed low-quality bases (Phred-scaled base quality < 20) and removed adapters with Trim Galore v. 0.6.5 (stringency=3)^47^. We used CutAdapt v.4.2 to further filter reads (--nextseq-trim=20 --minimum-length=20 --pair-filter=any)^48^.To exclude potential contamination which a previous study shows can be a source of false genetic variation^49^, we used Kraken2 to taxonomically classify reads and remove reads that were not assigned to the *Mycobacterium* genus or that were assigned to a *Mycobacterium* species other than *M. tuberculosis*^50^. We mapped reads with bwa v. 0.7.15 (*bwa* mem)^43^ to the H37Rv reference genome (NCBI Accession: NC_000962.3 [https://www.ncbi.nlm.nih.gov/nuccore/NC_000962.3]) and removed duplicates with sambamba^51^. We called variants with GATK 4.1 HaplotypeCaller^44^, setting sample ploidy to 1, and GenotypeGVCFs, including non-variant sites in output VCF files. We included variant sites with a minimum depth of 5X and a minimum variant quality score 20 and constructed consensus sequences with bcftools consensus^52^, excluding indels. We flagged SNPs in previously defined repetitive regions (PPE and PE-PGRS genes, phages, insertion sequences and repeats longer than 50 bp)^53^ and excluded these variants in figures and statistics except when otherwise noted. We identified sub-lineage and evidence of mixed infection with TBProfiler v.4.2.0^54,55^.

We constructed full-length consensus FASTA sequences from VCF files, setting missing genotypes to missing, and used SNP-sites to extract a multiple alignment of internal variant sites only^56^. We used the R package *ape* to measure pairwise differences between samples (*dist.dna,* pairwise.deletion=TRUE)^57^. We selected a best fit substitution model with ModelFinder^58^, implemented in IQ-TREE multicore version 2.2.0^59^, evaluating all models that included an ascertainment bias correction for the use of an alignment of SNPs only. We then fit a maximum likelihood tree with IQ-TREE, with 1000 ultrafast bootstrap replicates^59,60^ to visualize the location of household pairs in the context of study-wide variation.

We filtered variants that had coverage higher or lower than two standard deviations from the sample mean depth, reasoning that the extreme coverage was a result of incorrect mapping. We considered minority variants as positions with two or more alleles each supported by at least 5X coverage at the same position, at first, without filtering by minor allele frequency threshold. To examine the impact of filtering approach on the informativeness of minority variation, we applied increasingly conservative minor allele thresholds, from 0.05% to 50%. We quantified the number of per-sample minority variants; the number of shared minority variants between household members, defined as sharing the same minor allele call at the same position; and the number of shared minority variants between epidemiologically unrelated pairs.

Following variant identification, all analyses were conducted in R version 4.2.2. All analysis scripts are available on GitHub (https://github.com/ksw9/mtb-within-host).

### Role of the funding source

The study sponsor played no role in study design; in the collection, analysis, and interpretation of data; in the writing of the report; and in the decision to submit the paper for publication.

## Results

### M. tuberculosis variation observed in household transmission studies

To characterize the epidemiological information held in within-host *M. tuberculosis* variation present in routinely generated Illumina sequence data from cultured isolates, we reanalyzed sequence data from three previously published *M. tuberculosis* transmission studies for which whole genome sequencing data and epidemiological linkages were publicly available. Studies were from different epidemiological settings and included (a) a household transmission study in Vitória, Brazil^40^ (Colangeli et al.), a retrospective population-based study of pediatric tuberculosis in British Columbia, Canada^41^ (Guthrie et al.), and (c) a retrospective population-based study in Oxfordshire, England^11^ (Walker et al.). Study design, sampling design, and culture and sequencing methods differed across studies (Table 1).

As reported in the original studies, we observed limited fixed variation between *M. tuberculosis* consensus sequences from isolates collected within the same household or among isolates from patients with epidemiolocal linkages compared to randomly selected pairs of sequences from the same population (Fig. 1a). Consensus *M. tuberculosis* sequences from epidemiologically linked individuals were phylogenetic nearest neighbors for each study (Fig. 1b). However, genetic distances between consensus sequences often exceeded commonly used 5 and 12 SNP thresholds^10,11^ for classifying isolates as potentially linked in transmission, with 44.4% (20/45) of household pairs not meeting a 5-SNP threshold and 15.6% (7/45) of household pairs not meeting a 12-SNP threshold (Fig. 1a). Twenty-four percent (11/45) of isolate pairs from epidemiologically linked individuals were within a genetic distance of 2 SNPs or less, underscoring that genomic distances alone may be limited in their resolution.

**Figure 1.**
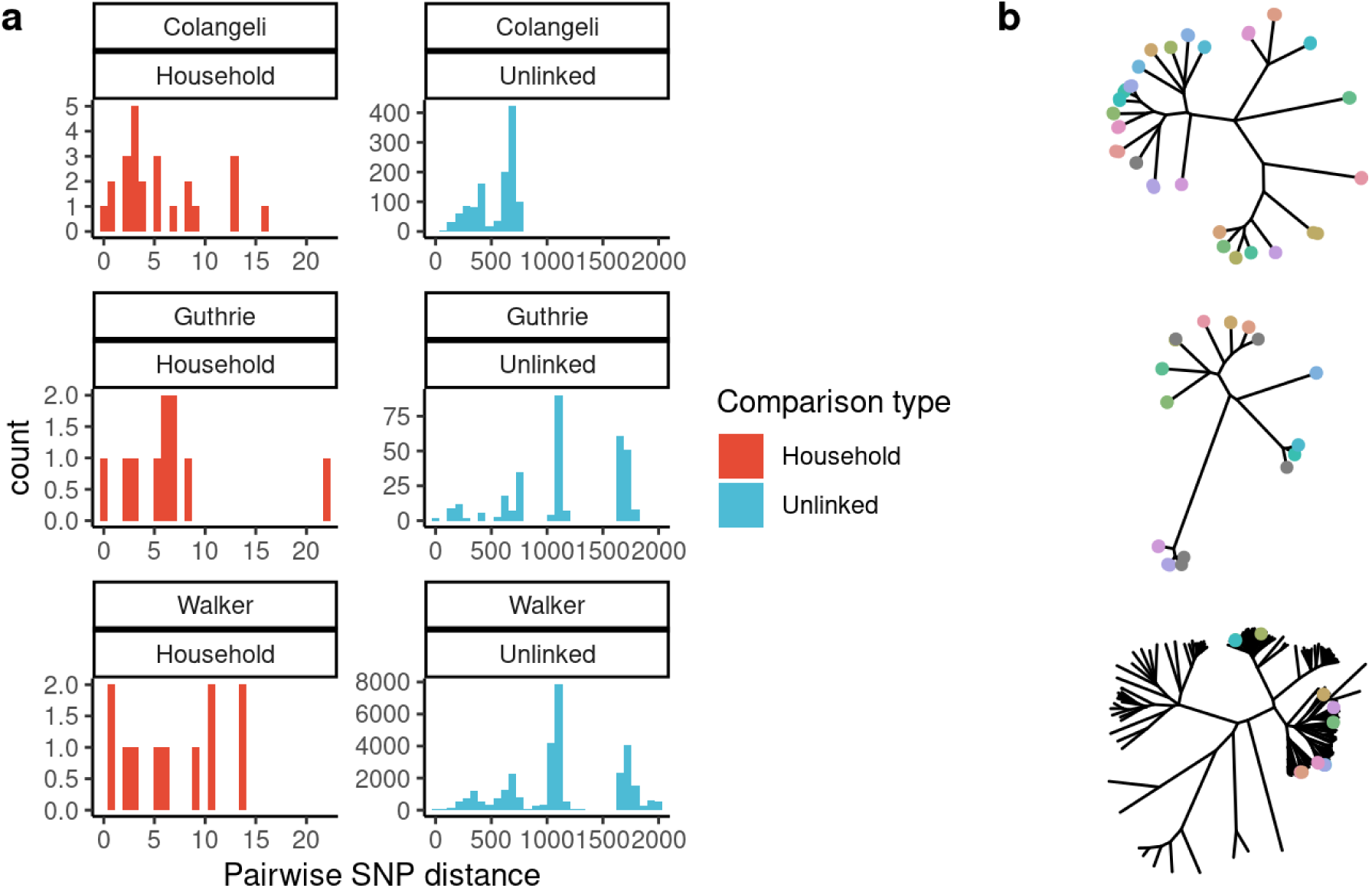
*M. tuberculosis* consensus genomes are closely related but are not always predictive of epidemiological linkage. (a) Histograms indicate pairwise genetic distances between *M. tuberculosis* consensus genomes, with facets indicating study and pairwise comparison type. (b) Phylogeny of consensus sequences for each study, with branch tips colored to indicate samples from a single household or with known epidemiologic links.

### Within-host variation observed in routine, culture-based M. tuberculosis sequencing data

We quantified minority variation within samples as the number of positions with a minor allele above a frequency of a range of threshold values, as we were interested in tradeoffs between sensitivity and specificity of variant detection. We detected limited, but measurable, minority variation above a 1% minor allele frequency threshold, with a disproportionate number of minority variants occurring within the PE/PPE genes (24.8%, 82.2%, and 80.1% of all minority variants, across the studies) (Fig. 2). We found substantial differences in minority variation detected across studies with the Colangeli et al. study (median: 160 minority variants, IQR:130-220) identifying a higher level of minority variation than both the Guthrie et al. study (median: 3, IQR:1-8; Wilcoxon test, p < 0.005) and the Walker et al. study (median: 2, IQR: 1-4, p < 0.005) (Table 3).

**Figure 2.**
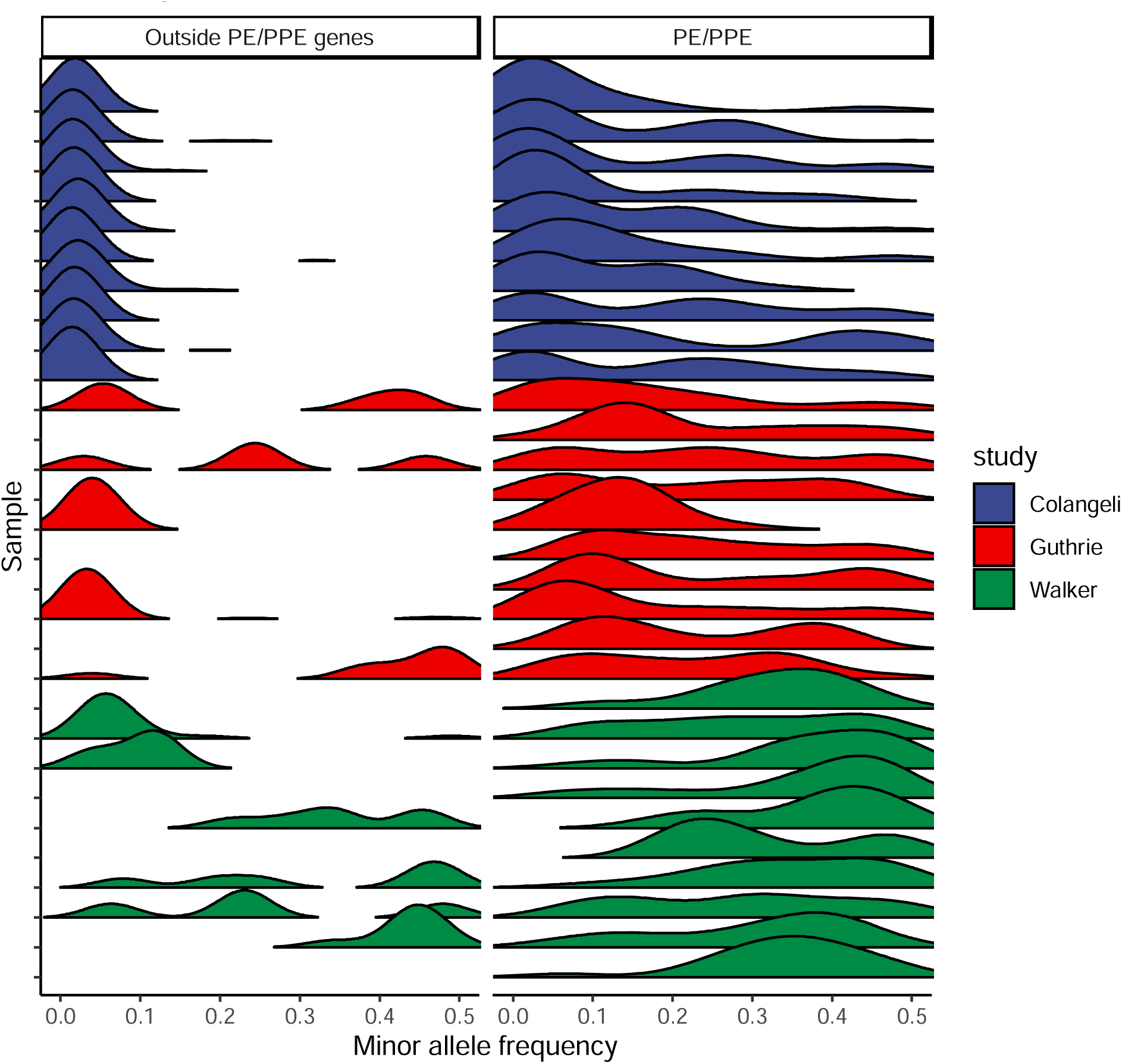
Limited *M. tuberculosis* within-host diversity is recovered with culture-based Illumina sequencing. Ridgeline plot of the minor allele frequency distribution for five randomly selected samples from each study, indicated by ridge color. Panels indicate genomic region: outside PE/PPE genes and within PE/PPE genes.

Most minority variants were in unique genomic locations and no minority variant was found in more than 5 samples in a single study (Fig. S1). About half of minority variants were predicted to be missense variants (50.0%; 964/1929) and only 1.3% (25/1929) minority variants were stop mutations, which would generate a truncated protein. However, the 5 most common minority variants across all three studies occurred in intergenic regions.

Median depth of coverage was significantly correlated with the total number of iSNVs detected outside the PE/PPE genes for the Walker et al. study, though no association was identified in the Colangeli et al. or Guthrie et al. studies (Fig. S2). Additionally, minor allele frequency was negatively correlated with site depth of coverage in the Colangeli et al. and the Walker et al. studies, but not Guthrie et al. (Fig. S2), potentially indicating that both culture method and sequencing depth were responsible for the observed differences in recovered variation (Table 1).

### Signatures of transmission in within-host M. tuberculosis variation

To test whether within-host variation could be used to identify potential transmission linkages, we quantified the number of shared minority variants passing quality, depth, and frequency thresholds between each pair of samples in each study. Isolates from household pairs shared more minority variants ≥1% frequency and outside of PE/PPE genes than did randomly selected pairs of isolates in all three studies (mean 98 shared minority variants vs. 10; 0.8 vs. 0.2; and 0.7 vs. 0.2, respectively; all p<0.001, Wilcoxon) (Table 2; Fig. 3). This effect rapidly declines as the definition of minority variant becomes more stringent (Fig. S3). In each study, the distribution of shared minority variants differed significantly between epidemiologically unlinked isolate pairs and epidemiologically linked pairs (Fig. 4a).

**Figure 3.**
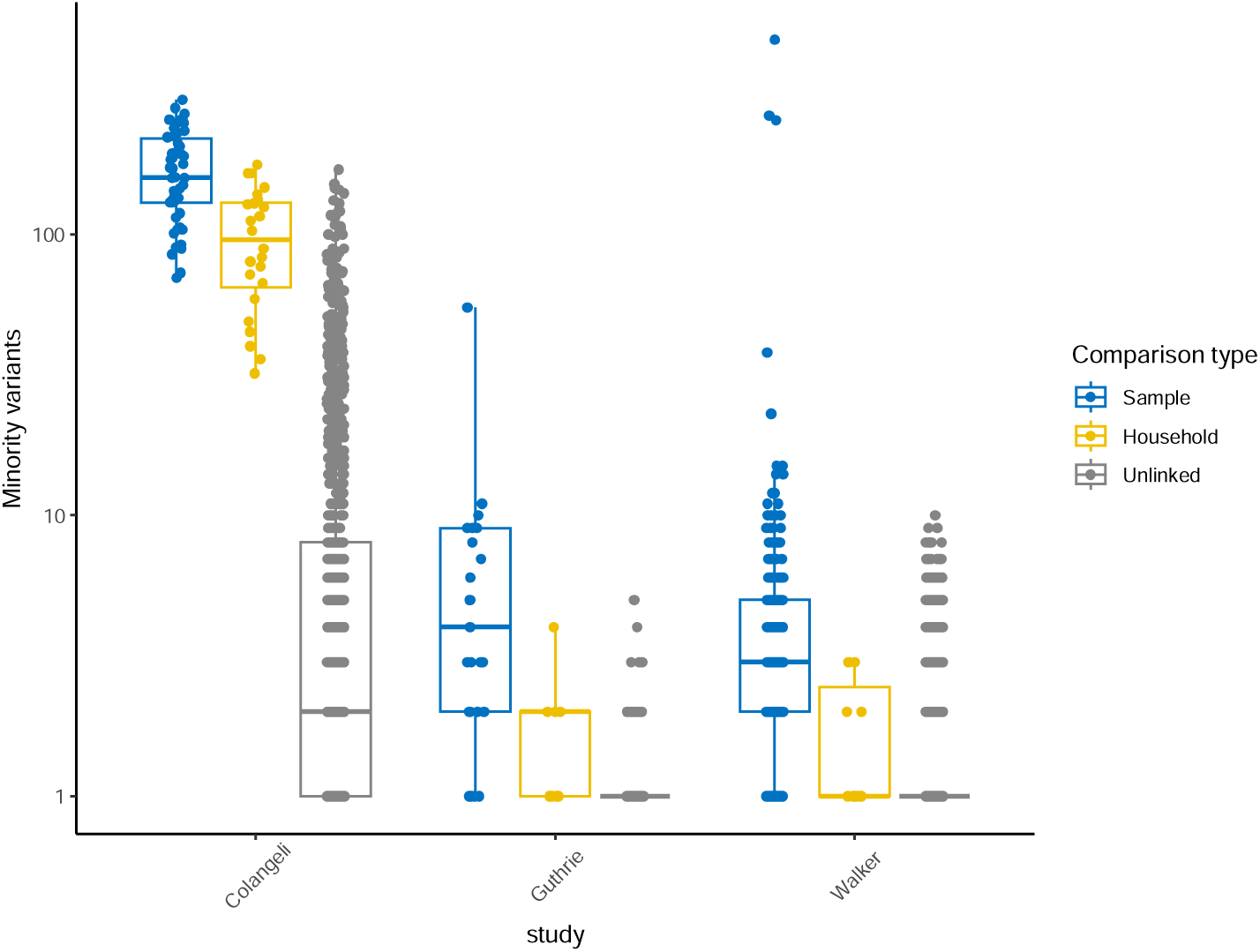
Pairwise shared variants above a 1% minor allele frequency. Boxplots of the number of high-quality shared minority variants between sample pairs in three previously published *M. tuberculosis* transmission studies (columns) with jittered points indicating pairwise observations. Colors indicate comparison type: sample, within-host minority variants; household, minority variants shared between household pairs; unlinked, minority variants shared between individuals in different households. Boxes indicate group interquartile ranges and center lines indicate group medians.

**Figure 4.**
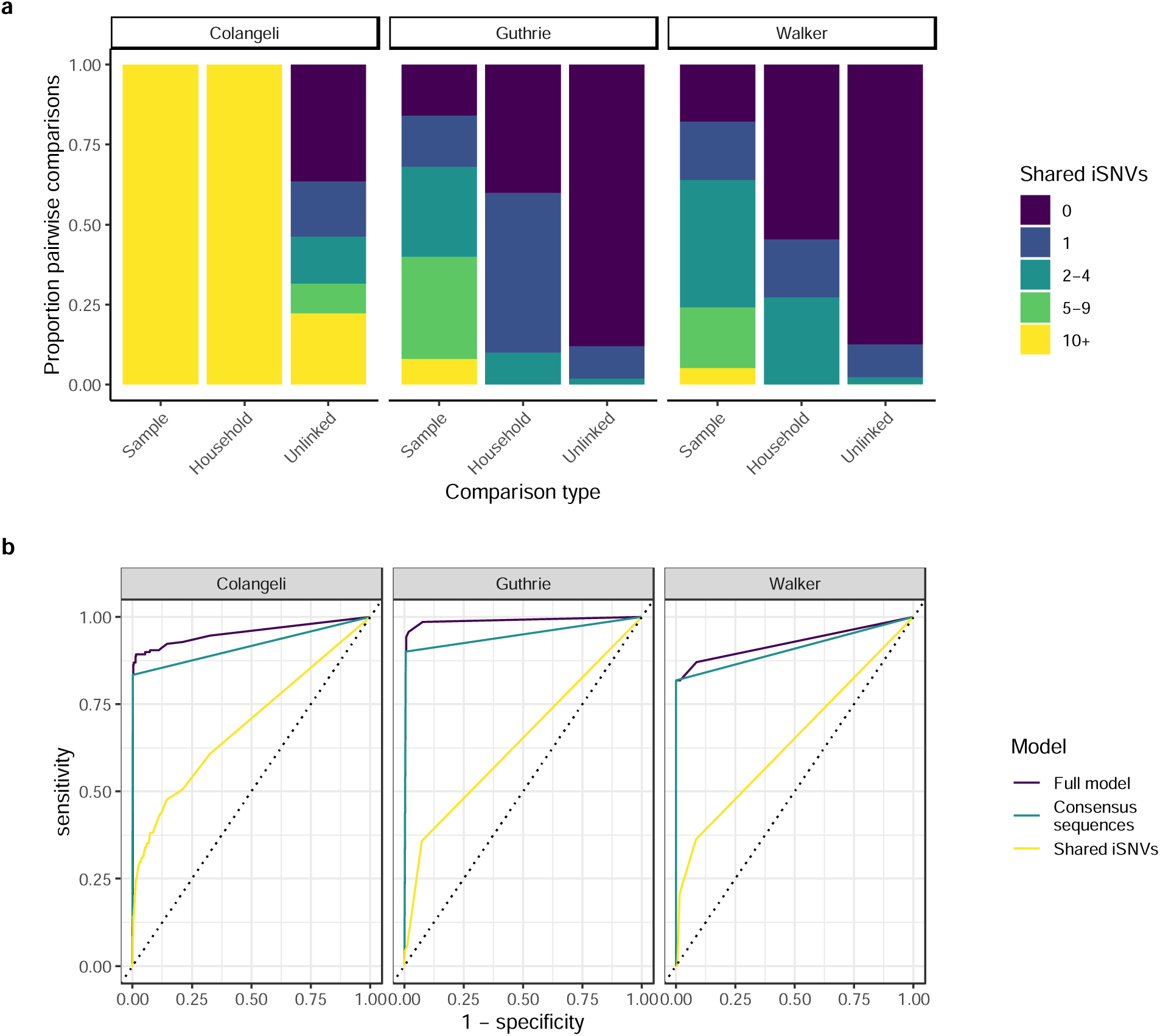
Shared minority variants contain information about household membership. (a) Stacked barplot showing the proportion of sample pairs across different levels of shared minority variants ≥ 1% minor allele frequency threshold. Panels indicate study. (b) ROC curves showing sensitivity (true positive rate) as a function 1 – specificity (true negative rate) for predicting household membership in general linear models that include both shared iSNVs and consensus sequence-based clusters (Full model), consensus sequence-based cluster only (Consensus sequences), and Shared iSNVs only (Shared iSNVs). All models include study as a predictor.

**Table 2.** Measured within-host *M. tuberculosis* variation. Per-sample and shared minority variants across pairwise comparisons with different epidemiological linkages, including minority variants ≥1% allele frequency, outside of the PE/PPE genes, and within an expected depth (defined in Methods).

| Comparison type | mean | median | lower | upper |
| --- | --- | --- | --- | --- |
| Colangeli |  |  |  |  |
| Sample | 170.00 | 160 | 130 | 220.0 |
| Household | 98.00 | 95 | 64 | 130.0 |
| Unlinked | 9.80 | 1 | 0 | 7.0 |
| Guthrie |  |  |  |  |
| Sample | 5.80 | 3 | 1 | 8.0 |
| Household | 0.80 | 1 | 0 | 1.0 |
| Unlinked | 0.15 | 0 | 0 | 0.0 |
| Walker |  |  |  |  |
| Sample | 7.10 | 2 | 1 | 4.0 |
| Household | 0.73 | 0 | 0 | 1.5 |
| Unlinked | 0.17 | 0 | 0 | 0.0 |

In a general linear model, shared within-host variation ≥1% frequency and outside of PE/PPE genes was significantly associated with household membership (OR: 1.51 [1.30,1.71] for one standard deviation increase in shared minority variants. Genomic clustering, based on a standard 12-SNP clustering distance thresholds, was also significantly associated with household membership (OR: 3,670 [1,160, 15,380]), with similar results when applying a 5-SNP clustering distance threshold. We measured the performance of general linear models in classifying household pairs versus unlinked pairs with receiver operator characteristic (ROC) curves. Including shared within-host variation improved the accuracy of predictions in all three studies as compared to a model without within-host variation (AUC: 0.95 *versus* 0.92, 0.99 *versus* 0.95, and 0.93 *versus* 0.91) (Fig. 4b). A model including within-host variation independently of consensus sequence-based clustering resulted in AUCs of 0.69, 0.64, and 0.64 for each study (Fig. 4b).

A major challenge in studies of pathogen variation and within-host variation, is distinguishing true biological variation from errors introduced through sampling, sequencing, and bioinformatic identification of variation in sequence data. To assess tradeoffs in sensitivity and specificity in minority variant identification, we applied a series of increasingly conservative minor allele frequency thresholds, filtering variants below a 0.05% to 50% frequency. Maximum AUC for predicting household membership was 0.998 (minor allele frequency threshold: 2%) for the Colangeli et al. study, 0.996 (threshold: 5%) for the Guthrie et al. study, and 0.94 (threshold: 5%) for the Walker et al. study (Fig. S4).

Among epidemiologically unlinked pairs, shared iSNVs declined significantly with increased genetic distance between samples across all studies (Fig. S5). For household pairs, we did not find a significant correlation between the genetic distance between isolate consensus sequences and number of shared minority variants in the Colangeli et al. and Walker et al. studies (Fig. S5), suggesting that this relationship may not be linear. While we did find a positive correlation between genetic distance and shared iSNVs for the Guthrie et al. study, this was due to a single pair with a genetic distance of greater than 20 SNPs.

Allele frequencies of shared minority variants ≥ 1% frequency located outside of PE/PPE genes were correlated between isolates from household pairs in Colangeli et al. (Pearson’s r=0.17, p<0.001) and Guthrie et al. (r=0.94, p <0.001), but not Walker et al. (Fig. S6). We predicted that sampling time might impact recovery of shared minority alleles because of changes in allele frequency between the time of sampling and time of transmission. Shared minority variation was negatively correlated with time between collection of isolates from household index cases and household members, though the association was not significant in the Colangeli et al. study, which reported time between sampling (Fig. S7).

## Discussion

To maximize the epidemiological information gleaned from the continuous evolution of *M. tuberculosis*, approaches to leverage biological variation more fully are needed. Here, we found that (1) within-host *M. tuberculosis* variation persists in sequence data from culture, (2) the magnitude of within-host variation varies between and within studies and is impacted by methodological choices, and (3) *M. tuberculosis* isolates from epidemiologically linked individuals share higher levels of variation than do unlinked individuals and shared within-host variation improves predictions of epidemiological linkage. Our results suggest that minority variation could contribute epidemiological information to transmission inferences, improving inferences from consensus sequences, and that alternative approaches to culture-based sequencing may further contribute to this observed epidemiological signal.

As sequencing has become more efficient and less expensive, pathogen genomic studies have begun to describe previously uncharacterized levels of minority variation within individual hosts and shared between transmission pairs. For example, *M. tuberculosis* within-host variation has been used to reveal an undetected superspreader event^25,26^ in a single large outbreak in the Canadian Arctic. In another study, Goig et al. observed minority variants that were shared between epidemiologically linked individuals, and one example of isolates from a four-person transmission cluster that all shared a minority variant at different allele frequencies^35^. The existence of shared minority variants suggests that multiple variants present in a donor’s infection persist through transmission and are maintained within the recipient through population changes and immune pressures. A similar observation has been made for other pathogens—shared within-host diversity of SARS-CoV-2 has been used to improve phylogenetic and transmission inferences in empirically collected and modeled sequence data^61–63^. Recently developed transmission inference approaches include pathogen within-host diversity to infer transmission events^64–67^, but have not yet been applied to *M. tuberculosis*, which is unique in its slow substitution rate and long and variable periods of latent infection. Future work is needed to develop automated, user-friendly pipelines for transmission and phylogenetic inference that include both fixed genomic differences and within-host variation.

Our findings that within-host diversity persists through culture and is impacted by methodological choices underscore the further work needed to optimize approaches for highly accurate identification of within-host variation. Each step of generating sequence data, including clinical sampling, sample preparation, sequencing, bioinformatic pipeline, may introduce a bottleneck and/or bias the variation recovered. For example, our observation that minority variants are concentrated in PE/PPE genes, highlights the need for testing whether long read sequencing or alternative mapping approaches can improve the accuracy of variant identification in this region^46^. Further, we found that increased sequencing coverage and, potentially, culture approach, detect higher levels of within-host variation.

A major challenge in pathogen genomics, including studies of within-host pathogen variation, is in distinguishing true biological variation from noise introduced by sequencing, bioinformatic, or other errors. There are significant trade-offs between sensitivity and specificity in variant identification; often, pathogen genomic approaches err on the side of specificity and impose conservative variant filters. Our findings here and previously^46^ suggest that for studying transmission linkages, including low frequency minority variants may improve predictions of transmission linkage. However, it is likely that some of the minority variants within individual samples and shared across samples are artefacts. For example, we found that some unlinked pairs of isolates share minority variants, potentially errors or true variants occurring at highly mutable sites (Fig. 3).

There are several limitations to our study. First, we conducted a re-analysis of previously published sequence data from clinical *M. tuberculosis* samples. We therefore do not have information about the true biological variation present within samples and cannot assess sensitivity and specificity of variants identified using alternative approaches. To measure performance of hybrid capture and other methods in recovering true within-host variation and the limit of detection of within-host variation, experiments that directly compare recovery of minority variants in known strain mixtures are required.

Second, we found that one study found substantially higher within-host variation than the others, likely reflecting large differences in study design and sample preparation (Table 1). The Colangeli et al. was a prospective study, and included three loops of culture for DNA extractions, while the Guthrie et al. and Walker et al. studies were retrospective and re-cultured isolates after frozen storage. This difference could also reflect higher population-wide *M. tuberculosis* diversity circulating in a higher-incidence setting. It is possible that other steps in *M. tuberculosis* sampling, sampling time (i.e. Fig. S5), culture, laboratory preparation, or sequencing influenced recovered within-host variation; if these steps were not reported, we were not able to include them in our models of within-host variation. For example, data on sequencing run, a potential source of false shared variation, was not available. Third, we considered household transmission pairs as our gold standard for transmission linkages. While the studies we included employed additional filters to exclude household pairs unlikely to be epidemiologically linked, it is possible that these pairs are misclassified. However, the impact of such misclassification would be to bias our results towards the null finding that shared minority variants are not more likely to be found in transmission pairs than unlinked pairs. Finally, we do not have access to sequencing replicates of the same sputum culture or biological replicates of the same sputum to quantify the concordance of minority variants across sequencing or biological replicates.

Our findings of within-host variation present in cultured *M. tuberculosis* samples suggests that within-host *M. tuberculosis* variation may be able to augment routine transmission inferences. More broadly, these finding suggests that assessing *M. tuberculosis* variation more broadly, including not only within-host variants, but also genome-wide variants and indels may yield more information and improve both transmission and phylogenetic inferences.

## Declaration of interest

The authors report no conflict of interest.

## Data Availability

We accessed publicly available data from 3 household transmission studies for which both raw sequence data and epidemiological linkages were publicly available: Colangeli et al. (2020), Vitoria, Brazil; Guthrie et al. (2018), British Columbia, Canada; and Walker et al. 2014, Oxfordshire, England (Table 1). Sequence data was available from the Sequence Read Archive (PRJNA475130, PRJNA413593, and PRJNA549270).

## Notes

### Competing Interest Statement

The authors have declared no competing interest.

### Funding Statement

This study was funded by NIAID, grant 5K01AI173385.

## References

1. Churchyard, G. et al. What We Know about Tuberculosis Transmission: An Overview. Journal of Infectious Diseases 216, S629–S635 (2017).

2. Auld, S. C. et al. Extensively drug-resistant tuberculosis in South Africa: Genomic evidence supporting transmission in communities. European Respiratory Journal 52, (2018).

3. Middelkoop, K. et al. Transmission of tuberculosis in a south African community with a high prevalence of HIV infection. Journal of Infectious Diseases 211, 53–61 (2015).

4. Andrews, J. R., Morrow, C., Walensky, R. P. & Wood, R. Integrating social contact and environmental data in evaluating tuberculosis transmission in a South African township. J Infect Dis 210, 597–603 (2014).

5. Yates, T. A. et al. The transmission of Mycobacterium tuberculosis in high burden settings. Lancet Infect Dis 16, 227–238 (2016).

6. Andrews, J. R., Morrow, C. & Wood, R. Modeling the role of public transportation in sustaining tuberculosis transmission in South Africa. Am J Epidemiol 177, 556–561 (2013).

7. Ypma, R. J. F., Altes, H. K., Van Soolingen, D., Wallinga, J. & Marijn Van Ballegooijen, W. A Sign of Superspreading in Tuberculosis: Highly Skewed Distribution of Genotypic Cluster Sizes. Source: Epidemiology 24, 395–400 (2013).

8. Gygli, S. M. et al. Prisons as ecological drivers of fitness-compensated multidrug-resistant Mycobacterium tuberculosis. Nat Med 27, 1171–1177 (2021).

9. Xu, Y. et al. High-resolution mapping of tuberculosis transmission: Whole genome sequencing and phylogenetic modelling of a cohort from Valencia Region, Spain. PLoS Med 16, (2019).

10. PHE. Tuberculosis in England: 2018 Presenting data to end of 2017. Public Health England **Version** 1., 173 (2018).

11. Walker, T. M. et al. Assessment of Mycobacterium tuberculosis transmission in Oxfordshire, UK, 2007-12, with whole pathogen genome sequences: An observational study. Lancet Respir Med 2, 285–292 (2014).

12. Bryant, J. M. et al. Inferring patient to patient transmission of Mycobacterium tuberculosis from whole genome sequencing data. BMC Infect Dis 13, 1–12 (2013).

13. Guerra-Assunção, J. et al. Large-scale whole genome sequencing of M. tuberculosis provides insights into transmission in a high prevalence area. Elife 4, 1–17 (2015).

14. Menardo, F., Duchêne, S., Brites, D. & Gagneux, S. The molecular clock of mycobacterium tuberculosis. PLoS Pathog 15, (2019).

15. Casali, N. et al. Evolution and transmission of drug-resistant tuberculosis in a Russian population. Nat Genet 46, 279–286 (2014).

16. Roetzer, A. et al. Whole Genome Sequencing versus Traditional Genotyping for Investigation of a Mycobacterium tuberculosis Outbreak: A Longitudinal Molecular Epidemiological Study. PLoS Med 10, (2013).

17. Yang, C., et al. Internal migration and transmission dynamics of tuberculosis in Shanghai, China: an epidemiological, spatial, genomic analysis. Lancet Infect Dis 18, 788–795 (2018).

18. Borges, V. et al. Nosocomial Outbreak of SARS-CoV-2 in a “Non-COVID-19” Hospital Ward: Virus Genome Sequencing as a Key Tool to Understand Cryptic Transmission. Viruses 13, (2021).

19. Cohen, T. et al. Mixed-Strain Mycobacterium tuberculosis Infections and the Implications for Tuberculosis Treatment and Control. Clin Microbiol Rev 25, 708–719 (2012).

20. Cohen, T. et al. Within-host heterogeneity of mycobacterium tuberculosis infection is associated with poor early treatment response: A prospective cohort study. Journal of Infectious Diseases 213, 1796–1799 (2016).

21. Pérez-Lago, L. et al. Revealing hidden clonal complexity in Mycobacterium tuberculosis infection by qualitative and quantitative improvement of sampling. Clinical Microbiology and Infection 21, 147.e1–147.e7 (2015).

22. Lieberman, T. D. et al. Genomic diversity in autopsy samples reveals within-host dissemination of HIV-associated Mycobacterium tuberculosis. Nat Med 22, 1470–1474 (2016).

23. Mann, B. C., Jacobson, K. R., Ghebrekristos, Y., Warren, R. M. & Farhat, M. R. Assessment and validation of enrichment and target capture approaches to improve Mycobacterium tuberculosis WGS from direct patient samples. J Clin Microbiol 61, (2023).

24. Séraphin, M. N. et al. Direct transmission of within-host Mycobacterium tuberculosis diversity to secondary cases can lead to variable between-host heterogeneity without de novo mutation: A genomic investigation. EBioMedicine 47, 293–300 (2019).

25. Lee, R. S., Proulx, J.-F. F., McIntosh, F., Behr, M. A. & Hanage, W. P. Previously undetected super-spreading of mycobacterium tuberculosis revealed by deep sequencing. Elife 9, (2020).

26. Martin, M. A., Lee, R. S., Cowley, L. A., Gardy, J. L. & Hanage, W. P. Within-host mycobacterium tuberculosis diversity and its utility for inferences of transmission. Microb Genom 4, (2018).

27. Zetola, N. M. et al. Mixed Mycobacterium tuberculosis complex infections and false-negative results for rifampin resistance by genexpert MTB/RIF are associated with poor clinical outcomes. J Clin Microbiol 52, 2422–2429 (2014).

28. Meehan, C. J. et al. Whole genome sequencing of Mycobacterium tuberculosis: current standards and open issues. Nat Rev Microbiol 17, 533–545 (2019).

29. McNerney, R. et al. Removing the bottleneck in whole genome sequencing of Mycobacterium tuberculosis for rapid drug resistance analysis: a call to action. International Journal of Infectious Diseases vol. 56 130–135 Preprint at 10.1016/j.ijid.2016.11.422 (2017).

30. Martín, A., Herranz, M., Ruiz Serrano, M. J., Bouza, E. & García de Viedma, D. The clonal composition of Mycobacterium tuberculosis in clinical specimens could be modified by culture. Tuberculosis 90, 201–207 (2010).

31. Plazzotta, G., Cohen, T. & Colijn, C. Magnitude and sources of bias in the detection of mixed strain M. tuberculosis infection. J Theor Biol 368, 67–73 (2015).

32. Walter, K. S. et al. Genomic variant-identification methods may alter mycobacterium tuberculosis transmission inferences. Microb Genom 6, 1–16 (2020).

33. Ates, L. S. New insights into the mycobacterial PE and PPE proteins provide a framework for future research. Mol Microbiol 0–2 (2019) doi:10.1111/mmi.14409.

34. Phelan, J. E. et al. Recombination in pe/ppe genes contributes to genetic variation in Mycobacterium tuberculosis lineages. BMC Genomics 17, 1–12 (2016).

35. Goig, G. A. et al. Whole-genome sequencing of Mycobacterium tuberculosis directly from clinical samples for high-resolution genomic epidemiology and drug resistance surveillance: an observational study. Lancet Microbe 1, e175–e183 (2020).

36. Brown, A. C. et al. Rapid whole-genome sequencing of mycobacterium tuberculosis isolates directly from clinical samples. J Clin Microbiol 53, 2230–2237 (2015).

37. Votintseva, A. A. et al. Same-day diagnostic and surveillance data for tuberculosis via whole-genome sequencing of direct respiratory samples. J Clin Microbiol 55, 1285–1298 (2017).

38. Nimmo, C. et al. Whole genome sequencing Mycobacterium tuberculosis directly from sputum identifies more genetic diversity than sequencing from culture. BMC Genomics 20, 1–9 (2019).

39. Doyle, R. M. et al. Direct Whole-Genome Sequencing of Sputum Accurately Identifies Drug-Resistant Mycobacterium tuberculosis Faster than MGIT Culture Sequencing. J Clin Microbiol 56, 666–684 (2018).

40. Colangeli, R. et al. Mycobacterium tuberculosis progresses through two phases of latent infection in humans. Nat Commun 11, (2020).

41. Guthrie, J. L. et al. Genotyping and Whole-Genome Sequencing to Identify Tuberculosis Transmission to Pediatric Patients in British Columbia, Canada, 2005-2014. J Infect Dis 218, 1155–1163 (2018).

42. Walter, K. S. et al. The role of prisons in disseminating tuberculosis in Brazil: A genomic epidemiology study. Lancet Regional Health -Americas 9, 100186 (2022).

43. Li, H. & Durbin, R. Fast and accurate short read alignment with Burrows-Wheeler transform. Bioinformatics 25, 1754–1760 (2009).

44. Van der Auwera, G. A. & O’Connor, B. *Genomics in the cloud*L*: using Docker, GATK, and WDL in Terra*. *Genomics in the cloud*L*: using Docker*, GATK, and WDL in Terra (O’Reilly Media, 2020).

45. Van der Auwera, G. A. et al. From FastQ Data to High-Confidence Variant Calls: The Genome Analysis Toolkit Best Practices Pipeline. in Current Protocols in Bioinformatics vol. 43 11.10.1-11.10.33 (John Wiley & Sons, Inc., 2013).

46. Walter, K. S. et al. Genomic variant-identification methods may alter Mycobacterium tuberculosis transmission inferences. Microb Genom 6, (2020).

47. Krueger, F. Trim Galore. Preprint at (2019).

48. Martin, M. Cutadapt Removes Adapter Sequences From High-Throughput Sequencing Reads. EMBnet J 17, (2011).

49. Goig, G. A., Blanco, S., Garcia-Basteiro, A. L. & Comas, I. Contaminant DNA in bacterial sequencing experiments is a major source of false genetic variability. BMC Biol 18, (2020).

50. Wood, D. E. & Salzberg, S. L. Kraken: Ultrafast metagenomic sequence classification using exact alignments. Genome Biol 15, R46 (2014).

51. Tarasov, A., Vilella, A. J., Cuppen, E., Nijman, I. J. & Prins, P. Sambamba: fast processing of NGS alignment formats. Bioinformatics 31, 2032–2034 (2015).

52. Danecek, P. et al. Twelve years of SAMtools and BCFtools. Gigascience 10, 1–4 (2021).

53. Brites, D. et al. A new phylogenetic framework for the animal-adapted mycobacterium tuberculosis complex. Front Microbiol 9, 2820 (2018).

54. Phelan, J. E. et al. Integrating informatics tools and portable sequencing technology for rapid detection of resistance to anti-tuberculous drugs. Genome Med 11, 41 (2019).

55. Coll, F. et al. Rapid determination of anti-tuberculosis drug resistance from whole-genome sequences. Genome Med 7, (2015).

56. Page, A. J. et al. SNP-sites: rapid efficient extraction of SNPs from multi-FASTA alignments. Microb Genom 2, 1–5 (2016).

57. Paradis, E. & Schliep, K. Ape 5.0: An environment for modern phylogenetics and evolutionary analyses in R. Bioinformatics 35, 526–528 (2019).

58. Kalyaanamoorthy, S., Minh, B. Q., Wong, T. K. F., Von Haeseler, A. & Jermiin, L. S. modelfinder: fast model selection for accurate phylogenetic estimates. 14, (2017).

59. Minh, B. Q. et al. IQ-TREE 2: New Models and Efficient Methods for Phylogenetic Inference in the Genomic Era. Mol Biol Evol 37, 1530–1534 (2020).

60. Hoang, D. T., Chernomor, O., Von Haeseler, A., Minh, B. Q. & Vinh, L. S. UFBoot2: Improving the Ultrafast Bootstrap Approximation. Mol Biol Evol 35, 518–522 (2018).

61. Torres Ortiz, A., et al. Within-host diversity improves phylogenetic and transmission reconstruction of SARS-CoV-2 outbreaks. doi:10.7554/eLife.

62. Walter, K. S. et al. Challenges in Harnessing Shared Within-Host Severe Acute Respiratory Syndrome Coronavirus 2 Variation for Transmission Inference. Open Forum Infect Dis 10, (2023).

63. Siddle, K. J. et al. Transmission from vaccinated individuals in a large SARS-CoV-2 Delta variant outbreak. Cell 185, 485–492.e10 (2022).

64. De Maio, N., Wu, C. H. & Wilson, D. J. SCOTTI: Efficient Reconstruction of Transmission within Outbreaks with the Structured Coalescent. PLoS Comput Biol 12, e1005130 (2016).

65. De Maio, N., Worby, C. J., Wilson, D. J. & Stoesser, N. Bayesian reconstruction of transmission within outbreaks using genomic variants. PLoS Comput Biol 14, e1006117 (2018).

66. Wymant, C. et al. PHYLOSCANNER: Inferring transmission from within- and between-host pathogen genetic diversity. Mol Biol Evol 35, 719–733 (2018).

67. Worby, C. J., Lipsitch, M. & Hanage, W. P. Shared Genomic Variants: Identification of Transmission Routes Using Pathogen Deep-Sequence Data. Am J Epidemiol 186, 1209–1216 (2017).

