## Supplemental Information for "Signatures of transmission in within-host *M. tuberculosis* variation"

**Supplementary Figures**

**Figure S1. Most observed minority variants occur at unique genomic locations.** Histogram of the count of isolate pairs with a minority variant at a specific genomic position. Facets indicate predicted mutational effect and study. Intergenic regions, missense, and synonymous variants are included, as they comprise the majority of predicted effects of observed minority variants.

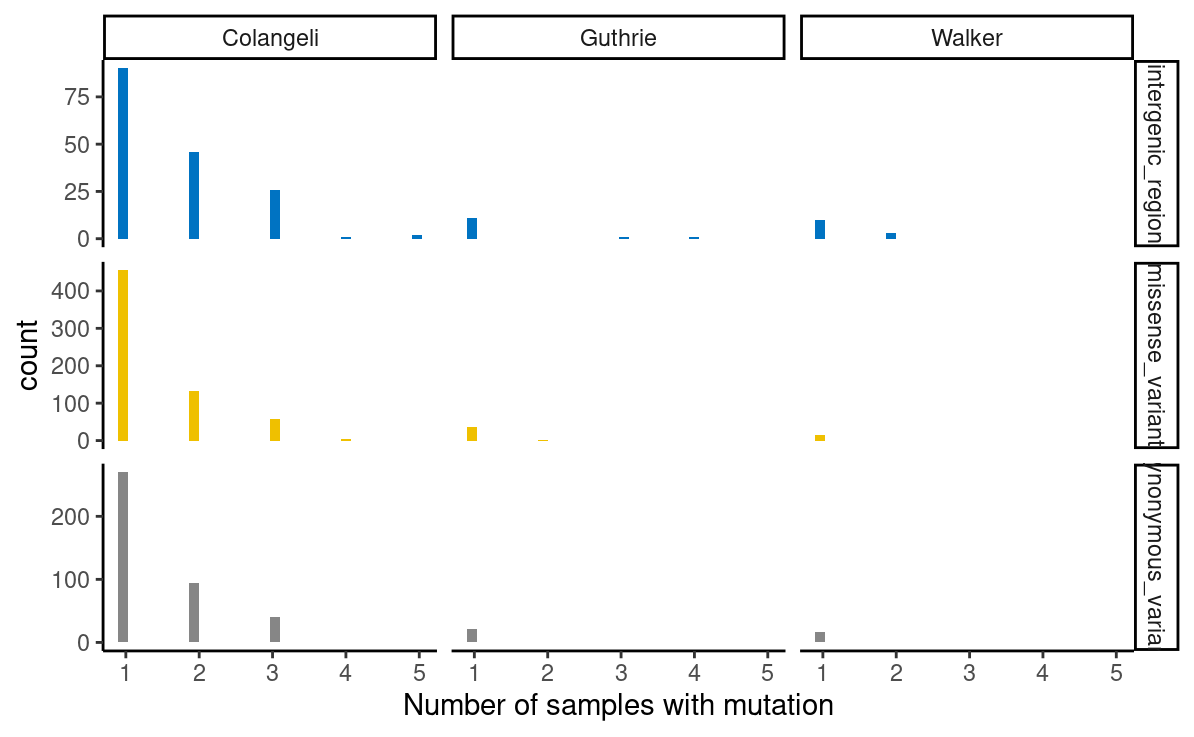

**Figure S2. Detection of minor alleles varies across studies.** Scatterplots of (a) the number of minority variants above a 1% minor allele frequency threshold as a function of sample median depth of coverage and (b) minor allele frequency as a function of per-site depth, faceted by study. Pearson’s correlation coefficient is reported for each study.

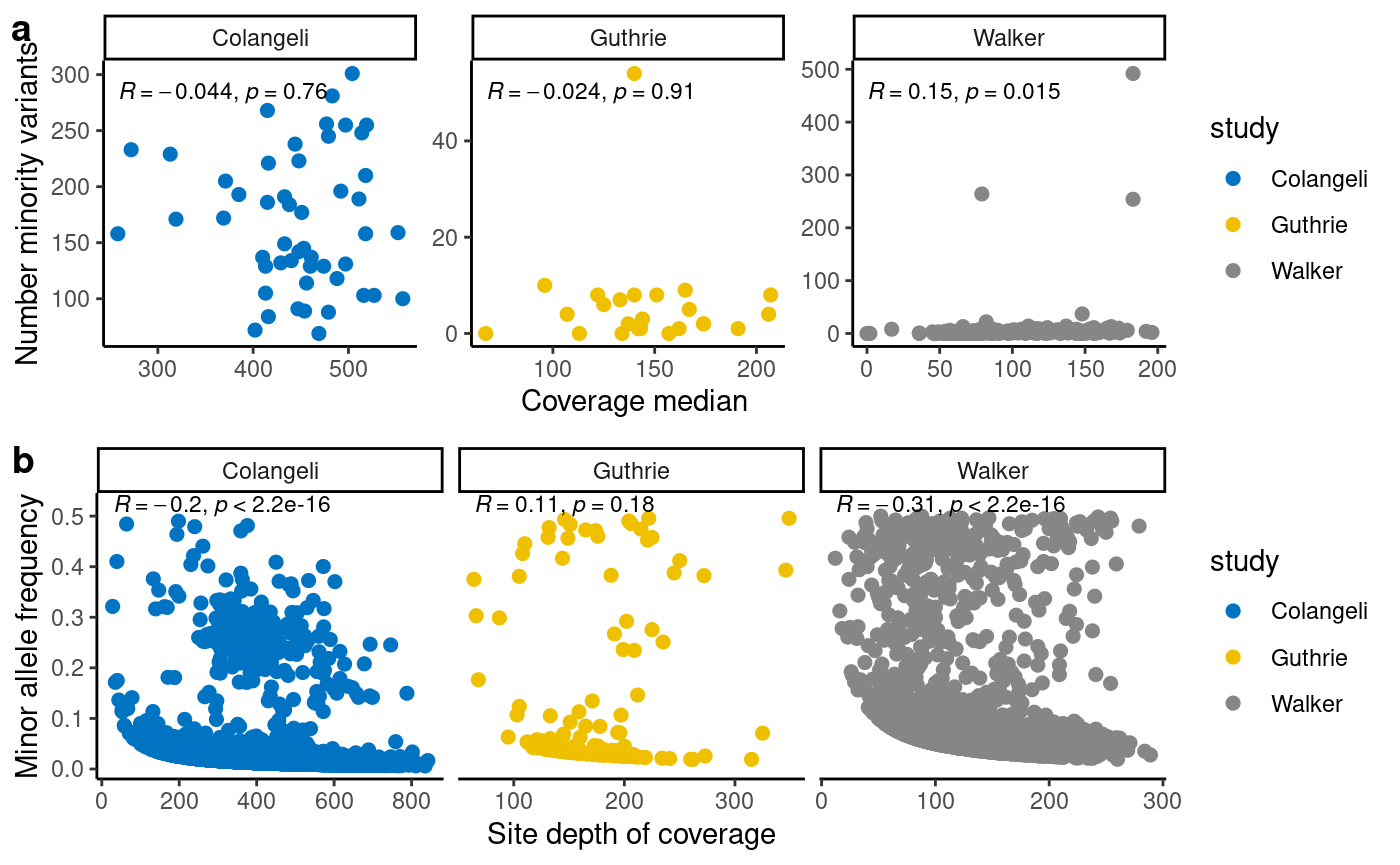

**Figure S3. Pairwise shared variants above alternate minor allele frequency thresholds.** Boxplots indicate the number of shared minority variants above 1, 5, and 10% minor allele frequency thresholds. Colors indicate comparison type: sample, within-host minority variants; household, minority variants shared between household pairs; outside household, minority variants shared between individuals in different households.

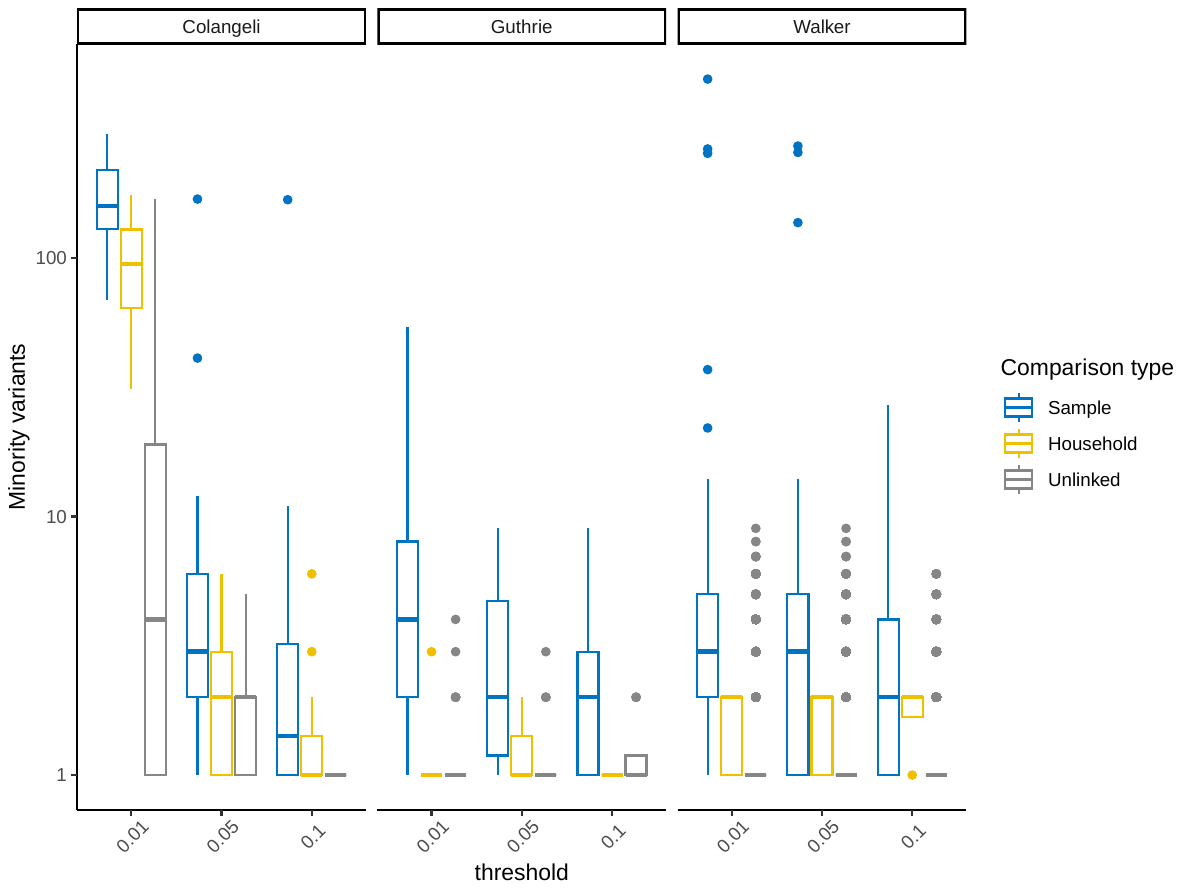

**Figure S4. Minor allele frequency threshold can alter the accuracy of predictions made with shared within-host variation.** ROC curves showing sensitivity (true positive rate) as a function 1 – specificity (true negative rate) for predicting household membership in general linear models that include both shared iSNVs and consensus sequence-based clusters when applying alternate minor allele frequency thresholds (color).

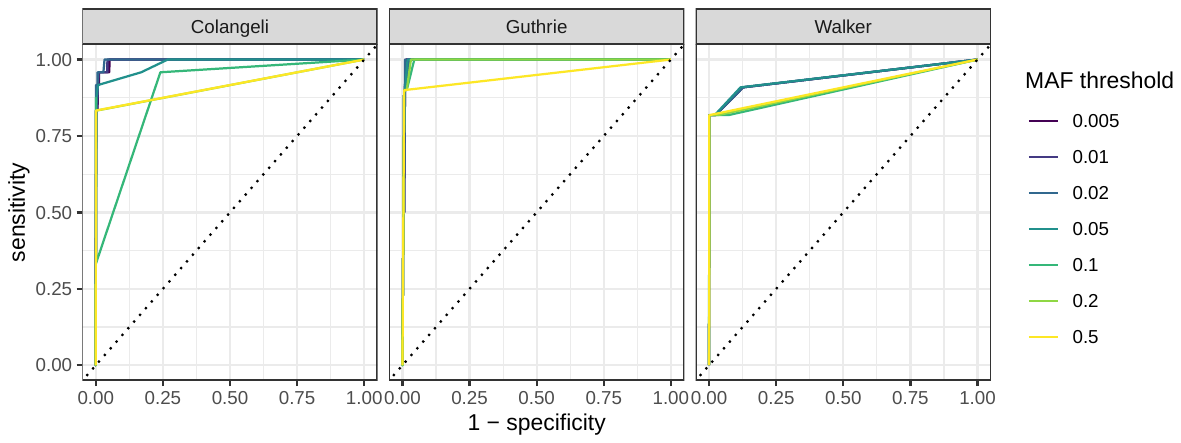

**Figure S5. Shared minority variants and distance between consensus sequences.** For each pair of isolates, the number of shared iSNVs versus the genetic distance between consensus sequences. Facets indicate study and comparison type and colors indicate study. Pearson’s correlation coefficient is reported for each study.

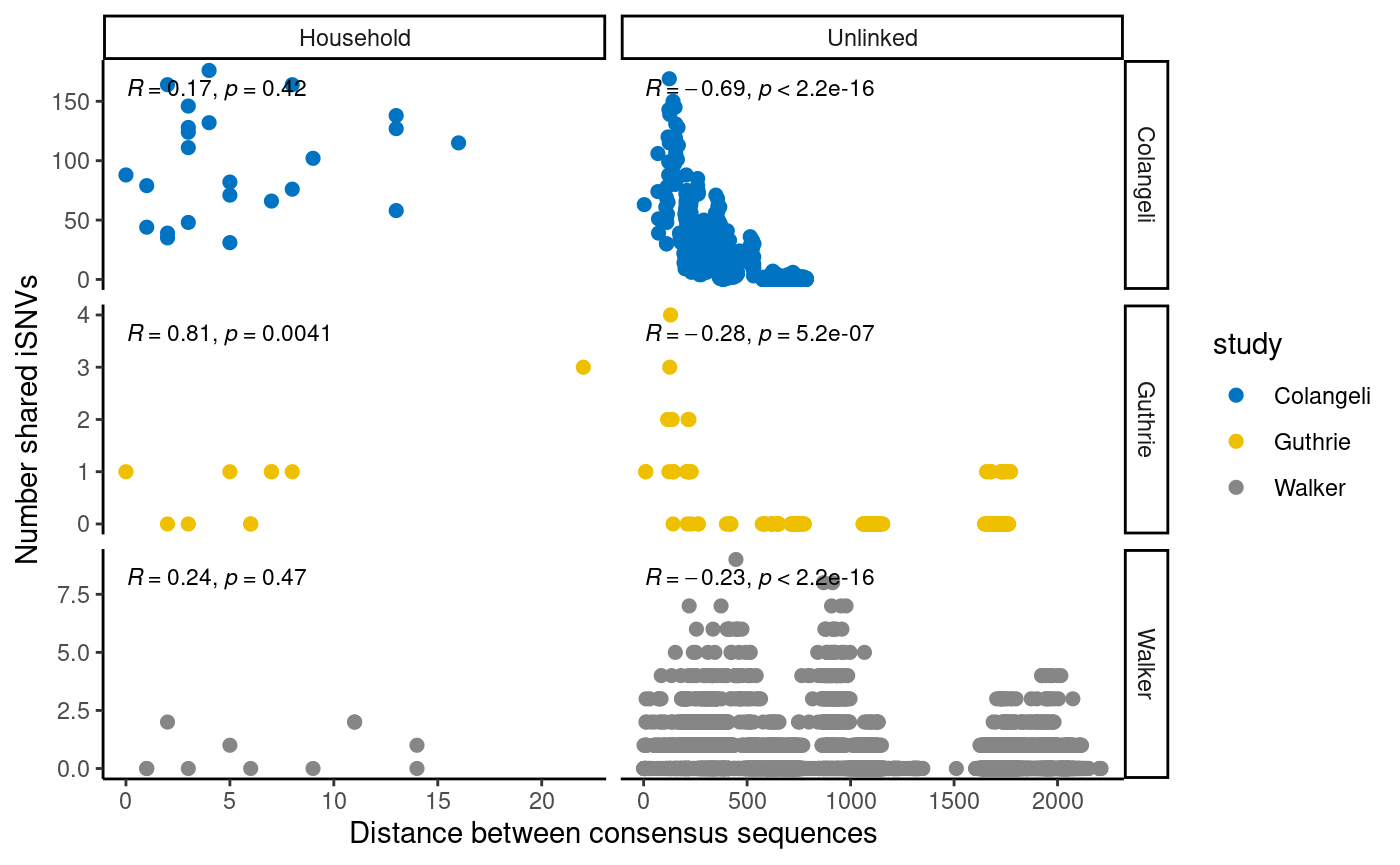

**Figure S6. Correlation between minor allele frequencies observed in shared iSNVs identified by GATK in transmission pairs.** Panel indicates study with Pearson’s correlation coefficient and p-value for each study.

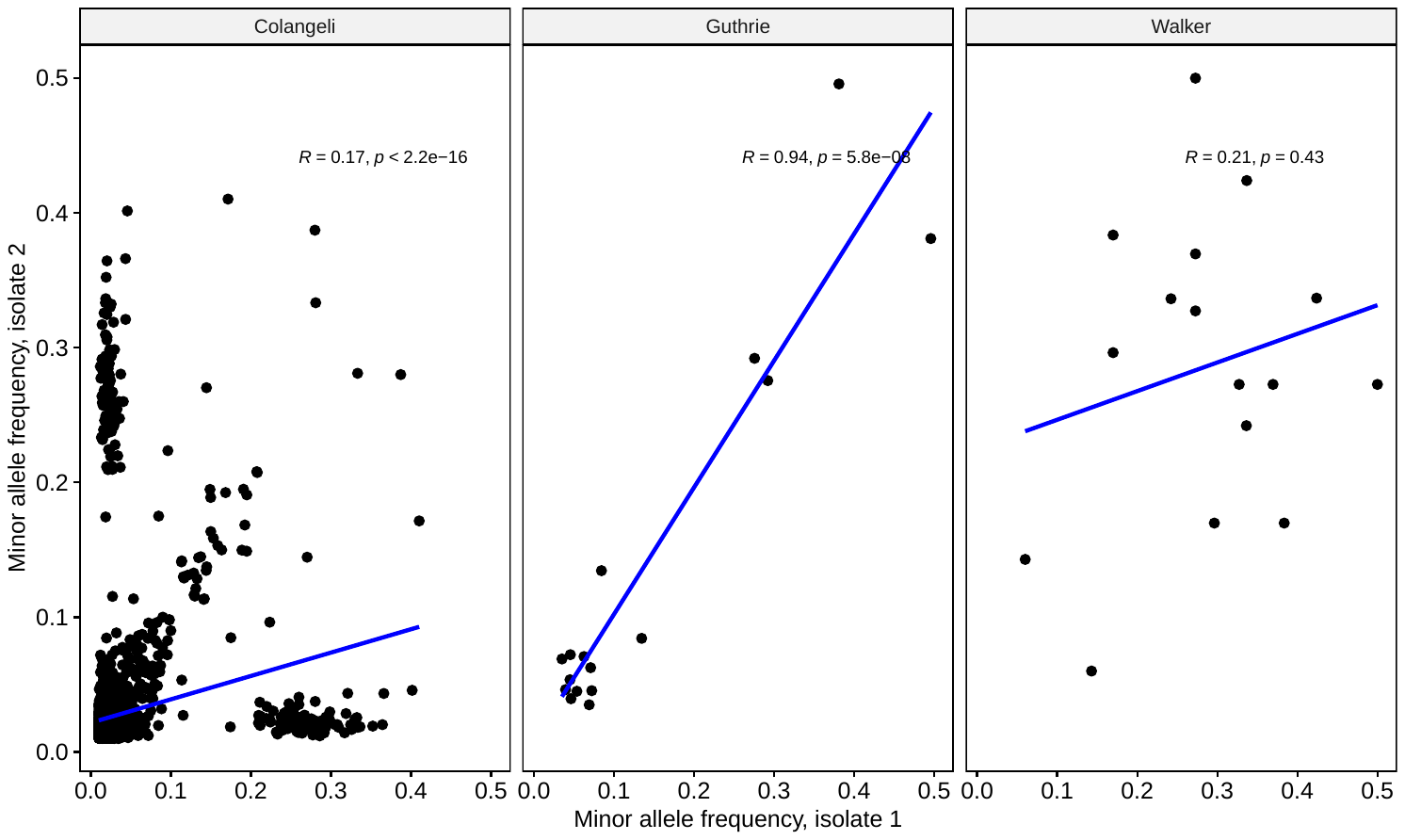

**Figure S7. Shared minority variants between household pairs declines within increased time between sample collection.** The number of shared minority variants plotted against months between index and recipient host diagnosis for each transmission pair from Colangeli et al.

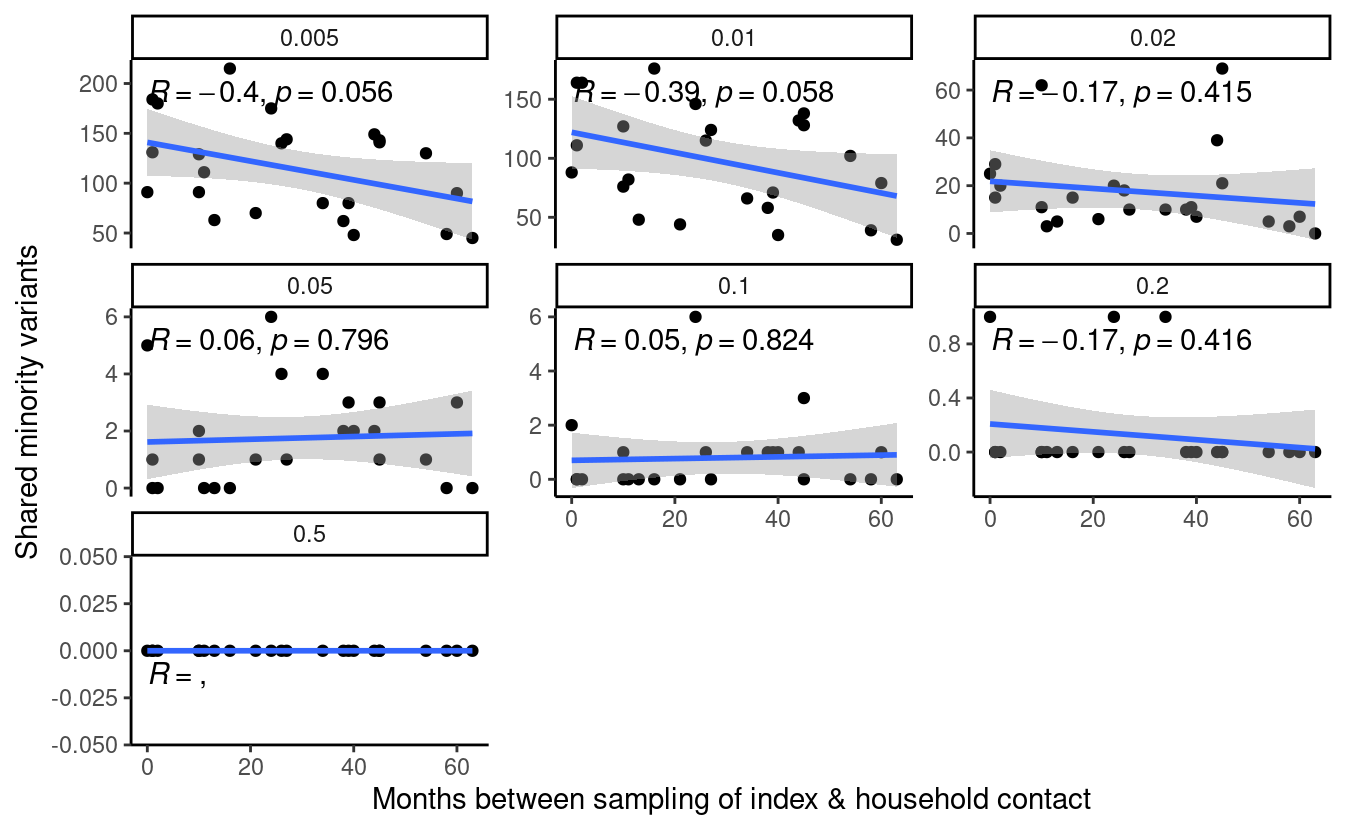
